# Observational Study of Haloperidol in Hospitalized Patients with Covid-19

**DOI:** 10.1101/2020.07.15.20150490

**Authors:** Nicolas Hoertel, Marina Sanchez Rico, Raphaël Vernet, Anne-Sophie Jannot, Antoine Neuraz, Carlos Blanco, Cédric Lemogne, Guillaume Airagnes, Nicolas Paris, Christel Daniel, Alexandre Gramfort, Guillaume Lemaitre, Mélodie Bernaux, Ali Bellamine, Nathanaël Beeker, Frédéric Limosin, On behalf of AP-HP / Universities / INSERM Covid-19 research collaboration and AP-HP Covid CDR Initiative

**Author notes:** Corresponding author Nicolas Hoertel, M.D., M.P.H., Ph.D., Corentin Celton Hospital, AP-HP.Centre, Paris University, 4 parvis Corentin Celton; 92130 Issy-les-Moulineaux, France, /. <u>Data Availability Statement</u> Data from the AP-HP Health Data Warehouse can be obtained at https://eds.aphp.fr//. <u>Disclaimer</u> The views and opinions expressed in this report are those of the authors and should not be construed to represent the views of any of the sponsoring organizations, agencies, or the US government. <u>Authorship</u> NH designed the study, performed statistical analyses, and wrote the first draft of the manuscript. MSR performed statistical analyses and critically revised the manuscript. RV contributed to statistical analyses and critically revised the manuscript for scientific content. FL, NB and ASJ contributed to study design and critically revised the manuscript for scientific content. NB, ASJ, AN, NP, CD, AG, GL, MB, and AB contributed to database build process. AN, CB, CL, GA, NP, CD, AG, GL, MB, and AB critically revised the manuscript for scientific content. <u>Funding source</u> This work did not receive any external funding. <u>Role of the funding source</u> None.

## Abstract

**Background:** Haloperidol, a widely used antipsychotic, has been suggested as potential effective treatment for Covid-19 on the grounds of its in-vitro antiviral effects against SARS-CoV-2.

**Methods:** We examined the association between haloperidol use and respiratory failure at AP-HP Greater Paris University hospitals. Data were obtained regarding all adult patients hospitalized with Covid-19 since the beginning of the epidemic. Study baseline was defined as the date of hospital admission. The primary endpoint was a composite of intubation or death and the secondary endpoint was discharge home among survivors in time-to-event analyses. We compared outcomes between patients who were exposed to haloperidol and those who were not, using a multivariable Cox regression model with inverse probability weighting according to the propensity score.

**Results:** Of the 13,279 hospitalized adult patients with positive Covid-19 RT-PCR test, 667 patients (5.0%) were excluded because of missing data. Of the remaining 12,612 patients, 104 (0.8%) were exposed to haloperidol. Over a mean follow-up of 20.8 days, the primary endpoint of respiratory failure respectively occurred in 27 patients (26.0%) exposed to haloperidol and 1,700 patients (13.6%) who were not. Among survivors, the secondary endpoint of discharge home occurred in 26 patients (32.1%) who received haloperidol and 6,110 patients (55.3%) who did not. In the main analysis, there were no significant associations between haloperidol use and the primary (HR, 1.09; 95% CI, 0.60 to 1.97, p=0.772) and secondary (HR, 0.88; 95% CI, 0.50 to 1.53, p=0.643) endpoints. Results were similar in multiple sensitivity analyses.

**Conclusion:** In this observational study involving patients with Covid-19 who had been admitted to the hospital, haloperidol use was not associated with risk of intubation or death, or with time to hospital discharge home. These results suggest that haloperidol is unlikely to have a clinical efficacy for Covid-19.

## 1. Introduction

The novel coronavirus SARS-CoV-2, the causative agent of coronavirus disease 2019 (Covid-19), has caused worldwide health, social and economic disruption. In the absence of a vaccine or antiviral medications with proven clinical efficacy,^1,2^ the search for an effective treatment for Covid-19 among all available medications is urgently needed.^2,3^

Based on recent advances in the knowledge of molecular details of SARS-CoV-2 infection,^2^ it has been suggested that two sets of pharmacological agents that show in-vitro antiviral activity should be prioritized in that search: the inhibitors of mRNA translation and the predicted regulators of the Sigma1 and Sigma2 receptors.^2^ Molecules that target Sigma receptors may reduce virus infectivity through different mechanisms, including lipid remodeling and endoplasmic reticulum stress reponse.^2,4^

Haloperidol, a butyrophenone-derivative antipsychotic widely used in the treatment of psychoses and delirium, has been suggested as potential effective treatment for Covid-19 on the grounds of its in-vitro antiviral effects against SARS-CoV-2 through Sigma receptor regulation.^2^ Short-term use of haloperidol is generally well tolerated,^5^ although side effects can occur, including extrapyramidal symptoms and QT interval prolongation.^6^

To our knowledge, no study has examined to date the efficacy of haloperidol for Covid-19 in clinical populations. Observational studies of patients with Covid-19 taking medications for other indications can help determine their efficacy for Covid-19 and decide which should be prioritized for randomized clinical trials, and minimize the risk for patients of being exposed to potentially harmful and ineffective treatments.

To this end, we took advantage of the continuously updated Assistance Publique-Hôpitaux de Paris (AP-HP) Health Data Warehouse, which includes all inpatient visits for Covid-19 to one of the 39 Greater Paris University hospitals since the beginning of the epidemic.

In this report, we examined the association between haloperidol use and respiratory failure among adult patients with Covid-19 who have been hospitalized in these medical centers. We hypothesized that haloperidol use would be associated with a lower risk of a composite endpoint of intubation or death, and with a shorter time from hospital admission to discharge home in time-to-event analyses that were adjusted for major predictors of respiratory failure and weighted according to propensity scores assessing the probability of haloperidol use.

## 2. Methods

### 2.1. Setting

We conducted this study at AP*-*HP, which comprises 39 hospitals, 23 of which are acute, 20 adult and 3 pediatric hospitals. We included all adults aged 18 years or over who have been admitted to the hospital from January 1^st^ until May 20^th^ with Covid-19, ascertained by a positive reverse-transcriptase–polymerase-chain-reaction (RT-PCR) test from analysis of nasopharyngeal or oropharyngeal swab specimens. The Institutional Review Board of the AP-HP clinical data warehouse approved this study (CSE-20-20_COVID19). All procedures related to this work adhered to the ethical standards of the relevant national and institutional committees on human experimentation and with the Helsinki Declaration of 1975, as revised in 2008.

### 2.2 Data sources

We used data from the AP-HP Health Data Warehouse (‘Entrepôt de Données de Santé (EDS)’). This warehouse contains all the clinical data available on all inpatient visits for Covid-19 to any of the 39 Greater Paris University hospitals. The data obtained included patients’ demographic characteristics, RT-PCR test results, medication administration data, past and current medication lists, past and current diagnoses, discharge disposition, ventilator use data, and death certificates.

### 2.3 Variables assessed

The following data were available for each patient at the time of the admission: sex; age; obesity (defined as having a body-mass index higher than 30 kg/m^2^ or an International Statistical Classification of Diseases and Related Health Problems (ICD-10) code for obesity (E66.0, E66.1, E66.2, E66.8, E66.9); self-reported smoking status; number of medical conditions associated with increased risk of severe SARS-CoV-2 infection,^7-11^ which were coded by practitioners based on ICD-10, including diabetes mellitus (E11), diseases of the circulatory system (I00-I99), diseases of the respiratory system (J00-J99), neoplasms (C00-C96), and diseases of the blood and blood-forming organs and certain disorders involving the immune mechanism (D5-D8); and any medication prescribed according to compassionate use or as part of clinical trials (e.g., hydroxychloroquine, azithromycin, remdesivir, tocilizumab, or sarilumab). To take into account possible confounding by indication bias for haloperidol, we included diagnosis of delirium, which was assessed using ICD-10 codes (F05 and R41.0), the number of current psychiatric disorders other than delirium (F00-F99 other than F05), the number of prescribed antipsychotics other than haloperidol; and the number of other prescribed psychotropic medications (i.e., antidepressants, benzodiazepines, Z-drugs, and mood stabilizers).

All medical notes and prescriptions are computerized in Greater Paris University hospitals. Medications and their mode of administration (i.e., dosage, frequency, date, condition of intake) were identified from medication administration data or scanned hand-written medical prescriptions, through two deep learning models based on BERT contextual embeddings,^12^ one for the medications and their mode of administration. The model was trained on the APmed corpus,^13^ a previously annotated dataset for this task. Extracted medications names were then normalized to the Anatomical Therapeutic Chemical (ATC) terminology using approximate string matching.

### 2.4 Exposure to haloperidol

Study baseline was defined as the date of hospital admission. Patients were considered to have been exposed to haloperidol if they were prescribed this medication at admission, ascertained by an ongoing medical prescription dated of less than 3 months before hospital admission, or if they were prescribed it during the follow-up period before the end of the hospitalization or intubation or death.

### 2.5 Endpoints

The primary endpoint was the time from study baseline to intubation or death. For patients who died after intubation, the timing of the primary endpoint was defined as the time of intubation. The secondary outcome was the time from study baseline to discharge home among survivors.

### 2.6 Statistical analysis

We calculated frequencies and means (± standard deviations (SD)) of each variable described above in patients exposed and not exposed to haloperidol and compared them using chi-square tests or Welch’s t-tests. Patients without an endpoint event had their data censored on May 20^th^, 2020.

To examine the association of haloperidol use with the primary composite endpoint of intubation or death and the secondary endpoint of discharge home among survivors, we performed Cox proportional-hazards regression models. Weighted Cox regression models were used when proportional hazards assumption was not met. To help account for the nonrandomized prescription of haloperidol and reduce the effects of confounding, the primary analysis used propensity score analysis with inverse probability weighting.^14,15^ The individual propensities for haloperidol prescription were estimated by a multivariable logistic regression model that included age, sex, obesity, smoking status, the number of medical conditions, any medication prescribed according to compassionate use or as part of a clinical trial, diagnosis of delirium, the number of other active psychiatric disorders, the number of prescribed antipsychotics outside haloperidol, and the number of other prescribed psychotropic medications. In the inverse-probability-weighted analysis, the predicted probabilities from the propensity-score model were used to calculate the stabilized inverse-probability-weighting weight.^14^ Associations between haloperidol use and the two outcomes were then estimated using multivariable Cox regression models. Kaplan-Meier curves were performed using the inverse-probability-weighting weights,^16^ and their 95% pointwise confidence intervals were estimated using the nonparametric bootstrap method.^17^

We conducted sensitivity analyses, including a multivariable Cox regression model comprising as covariates the same variables as the inverse-probability-weighted analysis, and a univariate Cox regression model in a matched analytic sample. For this latter analysis, we selected five controls for each exposed case, based on the same variables used for both the inverse-probability-weighted analysis and the multivariable Cox regression. To reduce the effects of confounding, optimal matching was used in order to obtain the smallest average absolute distance across all these characteristics between each exposed patient and its corresponding non-exposed matched controls.

Finally, within the group of patients exposed to haloperidol, we tested the association of daily dosage and duration of exposure (dichotomized into ‘prescription that began during the hospitalization’ and ‘prescription that started before admission’) with the two outcomes.

For all significant associations, we performed residual analyses to assess the fit of the data, check assumptions, including proportional hazards assumption, and examined the potential presence of outliers. To improve the quality of result reporting, we followed the recommendations of The Strengthening the Reporting of Observational Studies in Epidemiology (STROBE) Initiative.^18^ Statistical significance was fixed a priori at p<0.05. All analyses were conducted between May 21 and May 27 in R software version 2.4.3.

## 3. Results

### 3.1. Characteristics of the cohort

Of the 13,279 hospitalized adult patients with a positive Covid-19 RT-PCR test, a total of 667 patients (5.0%) were excluded because of missing data. Of the remaining 12,612 inpatients, 104 patients (0.8%) were exposed to haloperidol, at a mean dosage of 3.6 mg per day (SD=4.3; range: 0.1 mg to 20.0 mg), and this exposition started before the hospitalization in more than a third of them (35.6%, n=37) (**Figure 1**).

**Figure 1.**
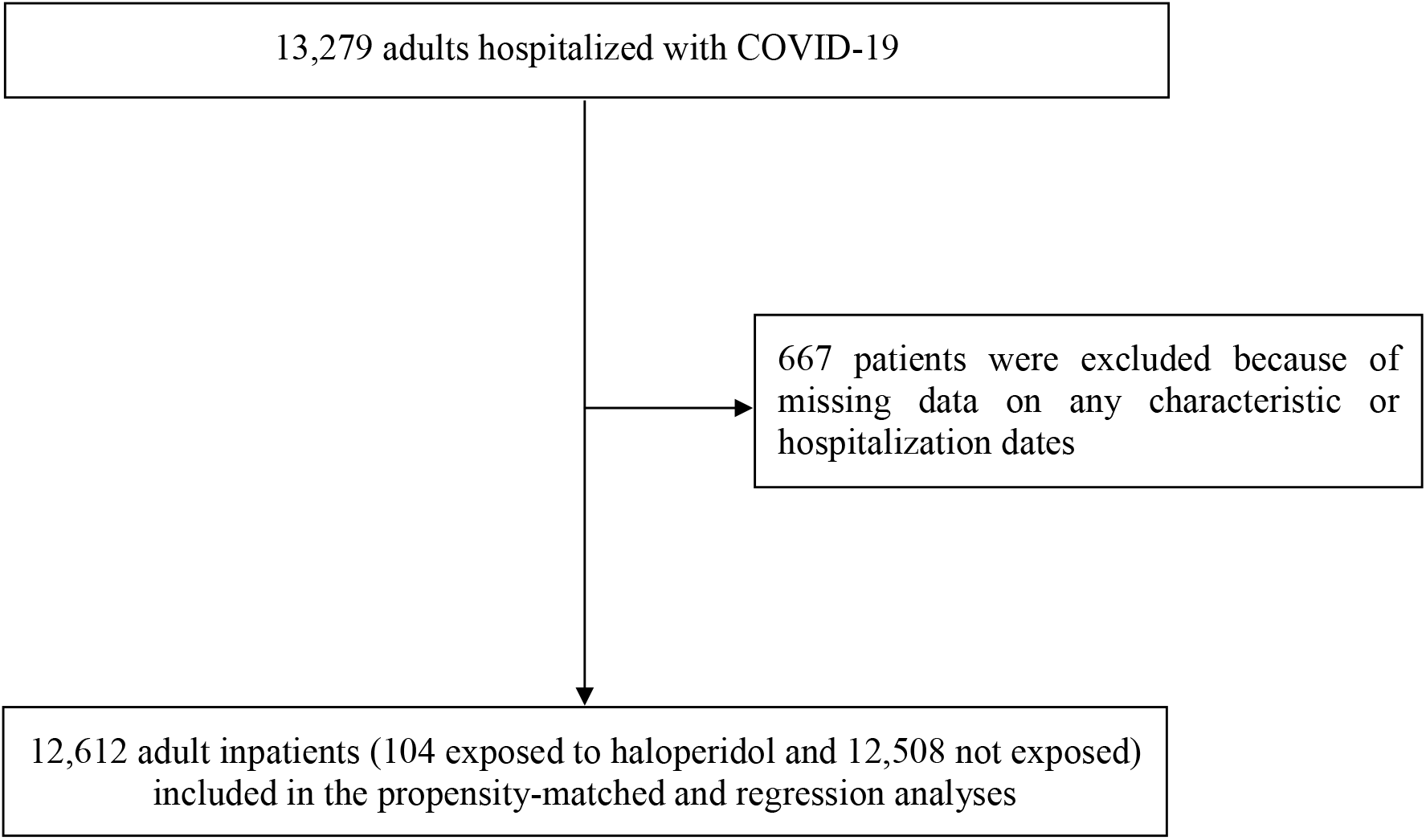
Study cohort.

Over a mean follow-up of 20.8 days (SD=28.0; median=7 days; range: from 1 day to 140 days), 1,727 patients (13.7%) had a primary end-point event and, among the 11,121 survivors, 6,136 patients (55.2%) were discharged home at the time of study end on May 20. Patients exposed to haloperidol had a mean follow-up of 27.5 days (SD=28.8; median=15 days; range: from 1 day to 127 days), while the non-exposed group had a mean follow-up of 20.8 days (SD = 28.0; median=7 days; range: from 1 day to 139 days).

The distribution of the patients’ characteristics according to haloperidol exposure is shown in **Table 1**. In the full sample, haloperidol exposure significantly differed according to age, smoking status, number of medical conditions, prevalence of delirium, number of other current psychiatric disorders, number of antipsychotics other than haloperidol, and number of other psychotropic medications, and the direction of associations indicated greater medical severity of people receiving haloperidol than those who did not. After applying the propensity score weights, these differences were substantially reduced but remained significant except for age, smoking status and number of medical conditions (**Table 1**).

**Table 1.** Characteristics of hospitalized patients with Covid-19 receiving or not receiving haloperidol in the matched and unmatched analytic samples.

|  | Exposed to<br>haloperidol<br>N = 104 | Not exposed to<br>haloperidol<br>N = 12,508 | Non-exposed<br>matched group<br>N = 520 | Exposed to<br>haloperidol<br>vs.<br>Not exposed to<br>haloperidol<br>(crude analysis) | Exposed to<br>haloperidol<br>vs.<br>Not exposed to<br>haloperidol<br>(analysis weighted<br>by inverse-probability-weighting weights) | Exposed to<br>Haloperidol<br>vs.<br>Non-exposed<br>matched group<br>(crude analysis<br>in the matched<br>analytic sample) |
| --- | --- | --- | --- | --- | --- | --- |
|  | N (%) | N (%) | N (%) | Chi-square test /<br>Welch's t-test (p-value) | Weighted Chi-square<br>test / Weighted<br>Welch's t-test (p-value) | Chi-square test /<br>Welch's t-test<br>(p-value) |
| *Characteristics* |  |  |  |  |  |  |
| Sex |  |  |  | 2.57 (0.109) | 0.16 (0.691) | 0.19 (0.665) |
| *Women* | 43 (41.3%) | 6,219 (49.7%) | 230 (44.2%) |  |  |  |
| *Men* | 61 (58.7%) | 6,289 (50.3%) | 290 (55.8%) |  |  |  |
| Age (years), mean (SD) | 69.9 (16.4) | 58.6 (20.0) | 72.1 (15.7) | -6.95 (<0.001*) | -0.63 (0.263) | -1.34 (0.184) |
| Smoker |  |  |  | 7.08 (0.008*) | 0.06 (0.812) | 0.01 (>0.99) |
| *Yes* | 18 (17.3%) | 1,153 (9.2%) | 87 (16.7%) |  |  |  |
| *No* | 86 (82.7%) | 11,355 (90.8%) | 433 (83.3%) |  |  |  |
| Obesity <sup>α</sup> |  |  |  | 1.44 (0.230) | 0.20 (0.658) | 0.08 (0.774) |
| *Yes* | 19 (18.3%) | 1,715 (13.7%) | 86 (16.5%) |  |  |  |
| *No* | 85 (81.7%) | 10,793 (86.3%) | 434 (83.5%) |  |  |  |
| Number of medical conditions |  |  |  | 12.36 (0.002*) | 2.46 (0.293) | 0.67 (0.716) |
| β |  |  |  |  |  |  |
| *0* | 58 (55.8%) | 8,932 (71.4%) | 287 (52.2%) |  |  |  |
| *1* | 10 (9.6%) | 808 (6.5%) | 39 (7.5%) |  |  |  |
| *2+* | 36 (34.6%) | 2,768 (22.1%) | 194 (37.3%) |  |  |  |
| Medication according to |  |  |  | 0.01 (>0.99) | 0.01 (0.916) | 0.01 (>0.99) |
| compassionate use or as part |  |  |  |  |  |  |
| of a clinical trial |  |  |  |  |  |  |
| *Yes* | 17 (16.3%) | 2,022 (16.2%) | 82 (15.8%) |  |  |  |
| *No* | 87 (83.7%) | 10,486 (83.8%) | 438 (84.2%) |  |  |  |
| Delirium <sup>£</sup> |  |  |  | 53.26 (<0.001*) | 25.70 (<0.001*) | 0.01 (>0.99) |
| *Yes* | 11 (10.6%) | 176 (1.41%) | 55 (10.6%) |  |  |  |
| *No* | 93 (89.4%) | 12332 (98.6%) | 465 (89.4%) |  |  |  |
| Number of current psychiatric |  |  |  | 94.29 (<0.001*) | 19.60 (<0.001*) | 0.01 (>0.99) |
| disorders <sup>¥</sup> |  |  |  |  |  |  |
| *0* | 78 (75.0%) | 11,918 (95.3%) | 390 (75.0%) |  |  |  |
| *1* | 15 (14.4%) | 384 (3.1%) | 75 (14.4%) |  |  |  |
| *2+* | 11 (10.6%) | 206 (1.7%) | 55 (10.6%) |  |  |  |
| Number of antipsychotics |  |  |  | 158.06 (<0.001*) | 23.91 (<0.001*) | 0.40 (0.817) |
| (other than haloperidol) |  |  |  |  |  |  |
| 0 | 78 (75.0%) | 12,120 (96.9%) | 403 (77.5%) |  |  |  |
| 1 | 19 (18.3%) | 302 (2.4%) | 82 (15.8%) |  |  |  |
| 2+ | 7 (6.7%) | 86 (0.7%) | 35 (6.73%) |  |  |  |
| Number of other psychotropic medications <sup>Ω</sup> |  |  |  | 366.94 (<0.001*) | 46.93 (<0.001*) | 0.28 (0.871) |
| 0 | 29 (27.9%) | 10,517 (84.1%) | 158 (30.4%) |  |  |  |
| 1 | 14 (13.5%) | 1,015 (8.1%) | 70 (13.5%) |  |  |  |
| 2+ | 61 (58.7%) | 976 (7.8%) | 292 (56.2%) |  |  |  |

<sup>α</sup> Defined as having a body-mass index higher than 30 kg/m<sup>2</sup> or based on ICD-10 codes (E66.0, E66.1, E66.2, E66.8, E66.9).
<sup>β</sup> Included diabetes mellitus (E11), diseases of the circulatory system (I00-I99), diseases of the respiratory system (J00-J99), neoplasms (C00-C96), and diseases of the blood and blood-forming organs and certain disorders involving the immune mechanism (D5-D8) based on ICD-10 codes.
<sup>£</sup> Assessed using ICD-10 codes (F05 and R41.0).
<sup>¥</sup> Assessed using ICD-10 codes (F00-F99) during the hospitalization.
<sup>Ω</sup> Included antidepressants, benzodiazepines, Z-drugs, and mood stabilizers (i.e., lithium and antiepileptic drugs).
\* p-value is significant (p<0.05).

In the matched analytic sample comprising 624 patients (i.e., 104 patients exposed to haloperidol and 520 patients from the matched group), there were no significant differences in any characteristics according to haloperidol exposure (**Table 1**).

### 3.2. Study endpoints

Respiratory failure respectively occurred in 27 patients (26.0%) who received haloperidol and 1,700 patients (13.6%) who did not (**Table 2**). There were no significant associations between haloperidol use and the composite primary endpoint both in the crude, unadjusted analysis (hazard ratio (HR), 1.38; 95% CI, 0.95 to 2.02; p=0.093) or in the propensity score weighted analysis (HR, 1.09; 95% CI, 0.60 to 1.97, p=0.772) (**Figure 2**; **Table 2**).

**Table 2.** Associations between haloperidol use and the endpoints of intubation or death and discharge home among survivors in the full sample and in the matched analytic sample of hospitalized patients with Covid-19.

| Analysis | Intubation or death | Discharge home among survivors |
| --- | --- | --- |
| ***Full sample*** |  |  |
| Number of events / Number of patients (%) | 1,727 / 12,612 (13.7%) | 6,136 / 11,121 (55.2%) |
| Haloperidol | 27 / 104 (26.0%) | 26 / 81 (32.1%) |
| No haloperidol | 1,700 / 12,508 (13.6%) | 6,110 / 11,040 (55.3%) |
| Crude analysis – hazard ratio (95% CI; p-value) | 1.38 (0.95 – 2.02; 0.093) | 0.33 (0.22 – 0.50; <0.001) |
| Multivariable analysis – hazard ratio (95% CI; p-value) | 0.77 (0.44 – 1.36; 0.369) | 1.06 (0.38 – 2.99; 0.917) |
| Propensity score analysis with inverse probability weighting – hazard ratio (95% CI; p-value) | 1.09 (0.60 – 1.97; 0.772) | 0.88 (0.50 – 1.53; 0.643) |
| ***Matched analytic sample*** |  |  |
| Number of events / Number of patients (%) | 178 / 624 (28.5%) | 179 / 464 (38.6%) |
| Haloperidol | 27 / 104 (26.0%) | 26 / 81 (32.1%) |
| No haloperidol | 151 / 520 (29.0%) | 153 / 382 (40.1%) |
| Crude analysis – hazard ratio (95% CI; p-value) | 0.76 (0.51 – 1.17; 0.223) | 0.84 (0.56 – 1.28; 0.420) |

**Figure 2.**
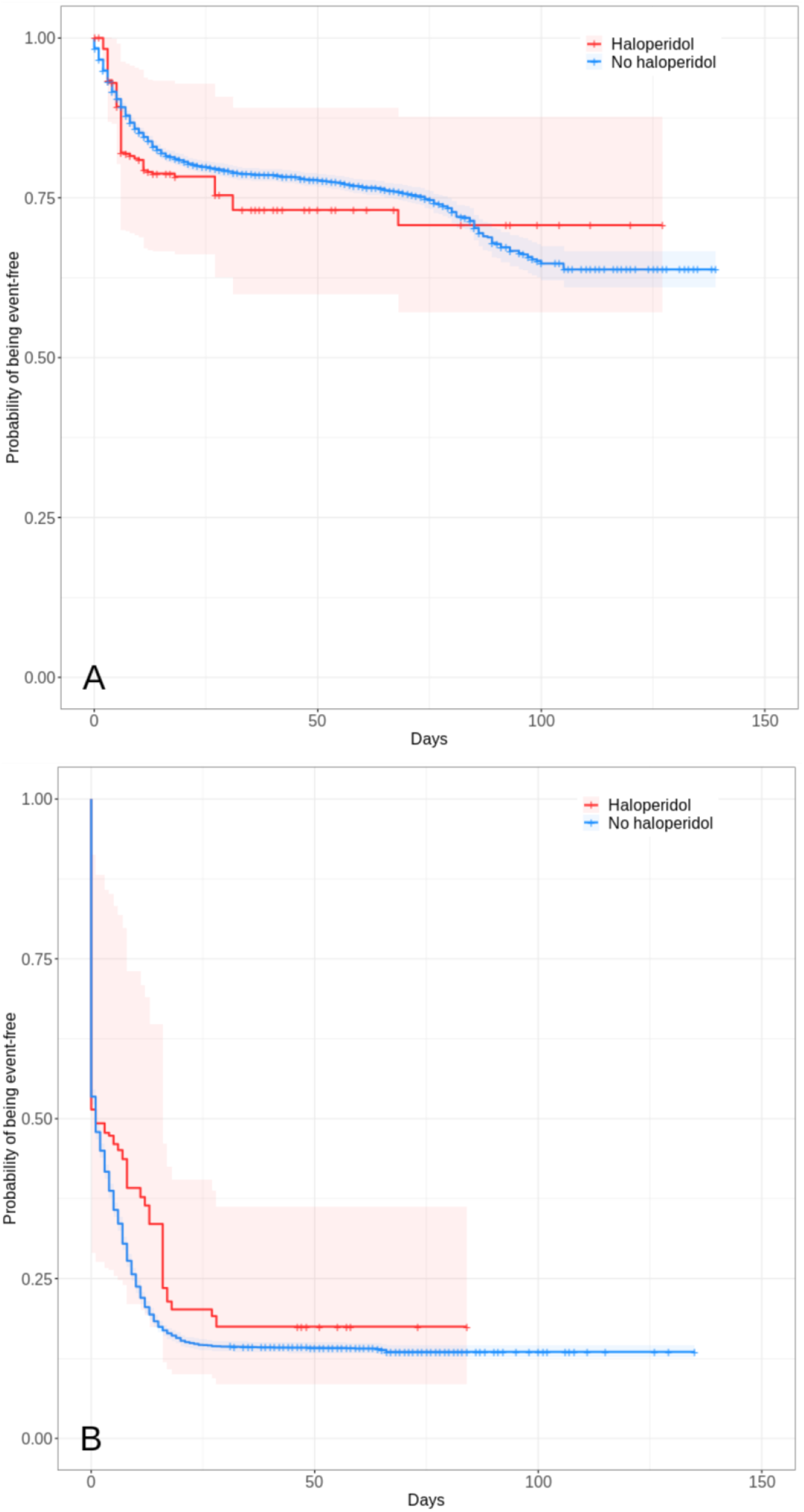
**Freedom from composite endpoint of intubation or death (N=12,612) (A) and from secondary endpoint of discharge home among survivors (N=11,121) (B) in the full sample of hospitalized patients with Covid-19 according to haloperidol exposure.^¥^** Note: The shaded areas represent pointwise 95% confidence intervals. ^¥^ Patients were considered to have been exposed to haloperidol if they were prescribed this medication at admission, ascertained by an ongoing medical prescription dated of less than 3 months before hospital admission, or if they were prescribed it during the follow-up period before the end of the hospitalization or intubation or death.

Among survivors, the secondary endpoint of discharge home occurred in 26 patients (32.1%) who were prescribed haloperidol and 6,110 patients (55.3%) who were not. Haloperidol use was significantly and negatively associated with the secondary endpoint in the crude, unadjusted analysis (HR, 0.33; 95% CI, 0.22 to 0.50, p<0.001), but not in the propensity score weighted analysis (HR, 0.88; 95% CI, 0.50 to 1.53, p=0.643) (**Figure 2**; **Table 2**).

Sensitivity analyses, including multivariable Cox regression models in the full sample and univariate Cox regression model in the matched analytic sample yielded similar non-significant results as the propensity score analyses for the two endpoints (**Figure 3**; **Table 2**). These results were unchanged when restricting the analyses to patients exposed to haloperidol only during the hospitalization for the two endpoints in the matched analytic sample analysis (HR, 0.75; 95% CI, 0.44 to 1.28, p=0.300 and HR, 0.75; 95% CI, 0.48 to 1.16, p=0.198, respectively) and for the primary endpoint in the propensity score weighted analysis (HR, 0.69; 95% CI, 0.37 to 1.26, p=0.222), but not for the secondary endpoint in that analysis, showing a significantly longer time to discharge home among survivors associated with haloperidol use (HR, 2.41; 95% CI, 1.75 to 3.31, p<0.001) (**Supplemental eFigures 1 and 2**). A post-hoc analysis indicated that in the full sample, we had 80% power to detect hazard ratios for haloperidol treatment of at least 1.15/0.86 for the primary endpoint and 1.11/0.90 for the secondary endpoint, while we had 80% power to detect hazard ratios of at least 1.61/0.58 for the primary endpoint and 1.65/0.59 for the secondary endpoint in the matched analytic sample.

**Figure 3.**
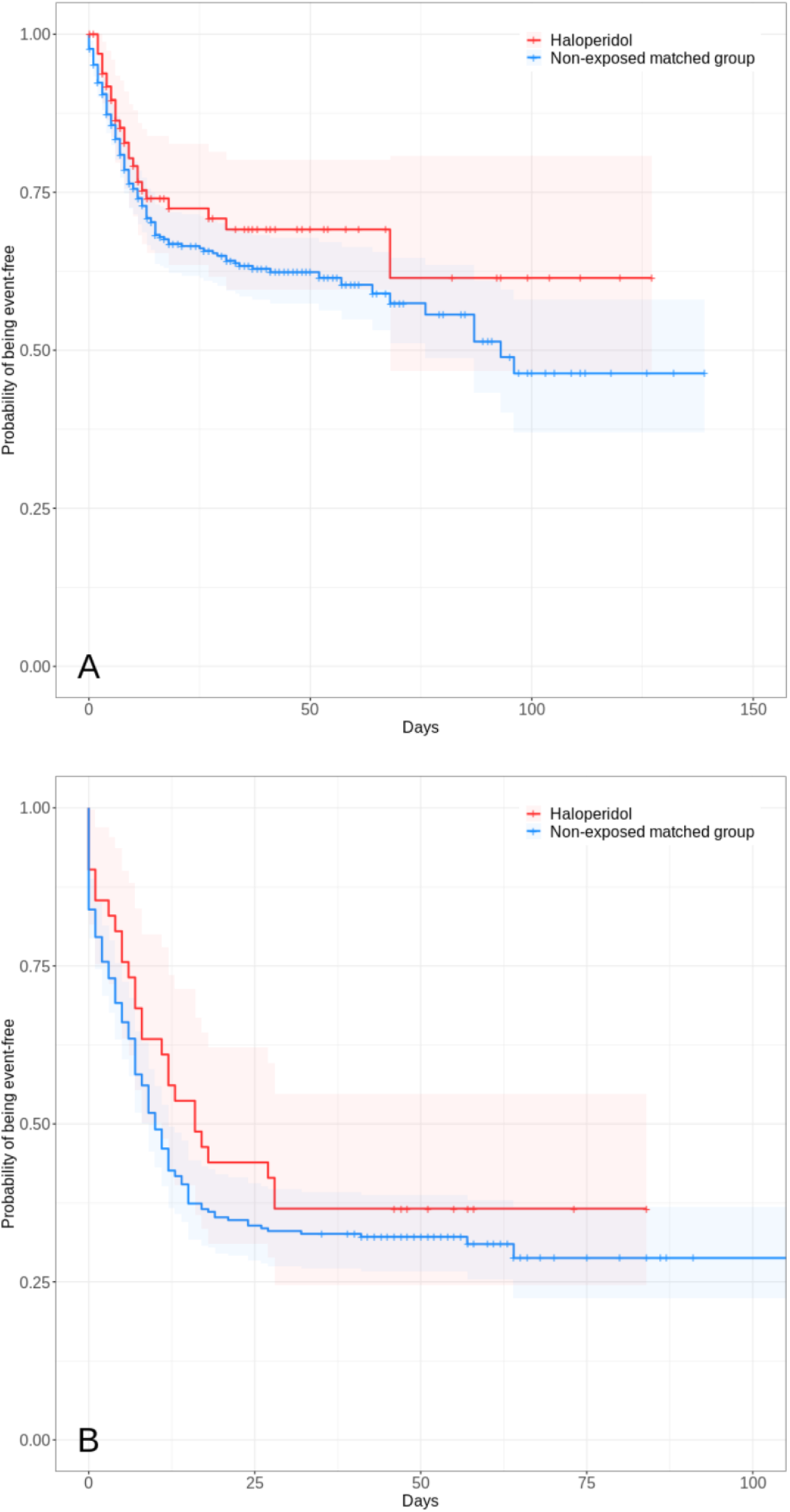
**Freedom from primary composite endpoint of intubation or death (N=624) (A) and from secondary endpoint of discharge home among survivors (N=464) (B) in the matched analytic sample of hospitalized patients with Covid-19 according to haloperidol exposure.^¥^** Note: The shaded areas represent pointwise 95% confidence intervals. ^¥^ Patients were considered to have been exposed to haloperidol if they were prescribed this medication at admission, ascertained by an ongoing medical prescription dated of less than 3 months before hospital admission, or if they were prescribed it during the follow-up period before the end of the hospitalization or intubation or death.

Finally, there were no significant associations of daily haloperidol dosage or duration of exposure within the group of patients exposed to haloperidol with the primary [HR (95% CI), 1.06 (0.95-1.18), p=0.297; HR (95% CI), 0.90 (0.41-1.94), p=0.779, respectively] or the secondary endpoint [HR (95% CI), 0.96 (0.84-1.08), p=0.480; HR (95% CI), 1.91 (0.53-6.90), p=0.324, respectively].

## Discussion

In this multicenter study involving a large number of inpatients with Covid-19, the risk of intubation or death and the time to discharge home among survivors were not significantly different between patients who were exposed to haloperidol and those who were not. Although these findings should be interpreted with caution due to the observational design, the relatively wide confidence intervals for estimates, and the fact that this is, to our knowledge, the first study examining the association of haloperidol use with respiratory failure in patients with Covid-19, they suggest that haloperidol is unlikely to have a clinically relevant efficacy for Covid-19 despite prior findings of in-vitro antiviral activity of this treatment against SARS-CoV-2.^2^

In the analyses, we tried to minimize the effects of confounding in several different ways. First, we used multivariable regression models with inverse probability weighting to minimize the effects of confounding by indication.^14,15^ We also performed sensitivity analyses, including a multivariable Cox regression model and a univariate Cox regression model in a matched analytic sample, which yielded similar results. Second, our analyses adjusted for potential confounders, including age, sex, obesity, smoking status, the number of medical conditions, any medication prescribed according to compassionate use or as part of a clinical trial, diagnosis of delirium, the number of other current psychiatric disorders, the number of prescribed antipsychotics other than haloperidol, and the number of other prescribed psychotropic medications. Although it is possible that some amount of unmeasured confounding remains, the consistency of results across the analyses gives support to our conclusion. Finally, the lack of significant associations of daily haloperidol dosage or duration of exposure with the two endpoints further supports our conclusion.

Additional limitations of our study include missing data for some variables (i.e., 5.0%) and potential for inaccuracies in the electronic health records, such as the possible lack of documentation of illnesses or medications, or the misidentification of treatments’ mode of administration (e.g., dosage, frequency), especially for hand-written medical prescriptions. However, results remained unchanged after using multiple imputation to account for missing data (available on request). Furthermore, patients under haloperidol were prescribed a relatively low dosage, i.e., 3.6 mg per day (SD=4.3), and its antiviral properties might be observable at higher dosages. However, we did not find any significant associations between dosage and the two endpoints. Finally, despite the multicenter design, our results may not be generalizable to other settings, e.g. outpatients, or regions.

In this observational study involving patients with Covid-19 who had been admitted to the hospital, haloperidol use was not associated with risk of intubation or death, or with time to hospital discharge home. These results suggest that haloperidol is unlikely to have a clinical efficacy for Covid-19.

## Data Availability

Data from the AP-HP Health Data Warehouse can be obtained at https://eds.aphp.fr//.

## Acknowledgments

The authors warmly thank the EDS APHP Covid consortium integrating the APHP Health Data Warehouse team as well as all the APHP staff and volunteers who contributed to the implementation of the EDS-Covid database and operating solutions for this database. Collaborators EDS APHP Covid consortium: Pierre-Yves ANCEL, Alain BAUCHET, Nathanaël BEEKER, Vincent BENOIT, Mélodie BERNAUX, Ali BELLAMINE, Romain BEY, Aurélie BOURMAUD, Stéphane BREANT, Anita BURGUN, Fabrice CARRAT, Charlotte CAUCHETEUX, Julien CHAMP, Sylvie CORMONT, Christel DANIEL, Julien DUBIEL, Catherine DUCLOAS, Loic ESTEVE, Marie FRANK, Nicolas GARCELON, Alexandre GRAMFORT, Nicolas GRIFFON, Olivier GRISEL, Martin GUILBAUD, Claire HASSEN-KHODJA, François HEMERY, Martin HILKA, Anne Sophie JANNOT, Jerome LAMBERT, Richard LAYESE, Judith LEBLANC, Léo LEBOUTER, Guillaume LEMAITRE, Damien LEPROVOST, Ivan LERNER, Kankoe LEVI SALLAH, Aurélien MAIRE, Marie-France MAMZER, Patricia MARTEL, Arthur MENSCH, Thomas MOREAU, Antoine NEURAZ, Nina ORLOVA, Nicolas PARIS, Bastien RANCE, Hélène RAVERA, Antoine ROZES, Elisa SALAMANCA, Arnaud SANDRIN, Patricia SERRE, Xavier TANNIER, Jean-Marc TRELUYER, Damien VAN GYSEL, Gaël VAROQUAUX, Jill Jen VIE, Maxime WACK, Perceval WAJSBURT, Demian WASSERMANN, Eric ZAPLETAL.

## References

1. Anderson RM, Heesterbeek H, Klinkenberg D, Hollingsworth TD. How will country-based mitigation measures influence the course of the COVID-19 epidemic? Lancet 2020; 395(10228): 931–4.

2. Gordon DE, Jang GM, Bouhaddou M, et al. A SARS-CoV-2 protein interaction map reveals targets for drug repurposing. Nature 2020:1–13.

3. Chevance A, Gourion D, Hoertel N, et al. Ensuring mental health care during the SARS-CoV-2 epidemic in France: a narrative review. L’encephale 2020.

4. Mitsuda T, Omi T, Tanimukai H, et al. Sigma-1Rs are upregulated via PERK/eIF2α/ATF4 pathway and execute protective function in ER stress. Biochemical and biophysical research communications 2011; 415(3): 519–25.

5. Zayed Y, Barbarawi M, Kheiri B, et al. Haloperidol for the management of delirium in adult intensive care unit patients: a systematic review and meta-analysis of randomized controlled trials. Journal of critical care 2019; 50: 280–6.

6. Xiong GL, Pinkhasov A, Mangal J, et al. QTc monitoring in adults with medical and psychiatric comorbidities: Expert consensus from the Association of Medicine and Psychiatry. Journal of Psychosomatic Research 2020: 110138.

7. Zhou F, Yu T, Du R, et al. Clinical course and risk factors for mortality of adult inpatients with COVID-19 in Wuhan, China: a retrospective cohort study. The Lancet 2020.

8. Ruan Q, Yang K, Wang W, Jiang L, Song J. Clinical predictors of mortality due to COVID-19 based on an analysis of data of 150 patients from Wuhan, China. Intensive care medicine 2020:1–3.

9. Du R-H, Liang L-R, Yang C-Q, et al. Predictors of mortality for patients with COVID-19 pneumonia caused by SARS-CoV-2: a prospective cohort study. European Respiratory Journal 2020; 55(5).

10. Hur K, Price CP, Gray EL, et al. Factors Associated With Intubation and Prolonged Intubation in Hospitalized Patients With COVID-19. Otolaryngology--Head and Neck Surgery.

11. Williamson E, Walker AJ, Bhaskaran KJ, et al. OpenSAFELY: factors associated with COVID-19-related hospital death in the linked electronic health records of 17 million adult NHS patients. medRxiv 2020.

12. Devlin J, Chang M-W, Lee K, Toutanova K. Bert: Pre-training of deep bidirectional transformers for language understanding. arXiv preprint 181004805 2018.

13. Jouffroy J, Feldman SF, Lerner I, Rance B, Neuraz A, Burgun A. MedExt: combining expert knowledge and deep learning for medication extraction from French clinical texts.

14. Robins JM, Hernan MA, Brumback B. Marginal structural models and causal inference in epidemiology. LWW; 2000.

15. Geleris J, Sun Y, Platt J, et al. Observational study of hydroxychloroquine in hospitalized patients with Covid-19. New England Journal of Medicine 2020.

16. Kaplan EL, Meier P. Nonparametric estimation from incomplete observations. Journal of the American statistical association 1958; 53(282): 457–81.

17. Efron B. Nonparametric standard errors and confidence intervals. canadian Journal of Statistics 1981; 9(2): 139–58.

18. Von Elm E, Altman DG, Egger M, Pocock SJ, Gøtzsche PC, Vandenbroucke JP. The Strengthening the Reporting of Observational Studies in Epidemiology (STROBE) statement: guidelines for reporting observational studies. Annals of internal medicine 2007; 147(8): 573–7.

